# Rare and common variant analyses of amyotrophic lateral sclerosis in the French-Canadian genome

**DOI:** 10.1101/2022.08.11.22278628

**Authors:** Jay P. Ross, Fulya Akçimen, Calwing Liao, Karina Kwan, Daniel E. Phillips, Zoe Schmilovich, Dan Spiegelman, Angela Genge, Nicolas Dupré, Patrick A. Dion, Sali M.K. Farhan, Guy A. Rouleau

**Author notes:** **Corresponding Author:** Dr. Guy A. Rouleau, MD, PhD, FRCPC, Department of Neurology and Neurosurgery, McGill University, Montréal, Québec, Canada, H3A 2B4.

## Abstract

The genetic etiology of ALS includes few rare, large-effect variants and potentially many common, small-effect variants per case. The genetic risk liability for ALS might require a threshold comprised of a certain amount of variants. Here, we tested the degree to which risk for ALS was affected by rare variants in ALS genes, polygenic risk score, or both. 335 ALS cases and 356 controls from Québec, Canada were concurrently tested by SNP-chip genotyping and targeted sequencing of known ALS genes. ALS GWAS summary statistics were used to estimate an ALS PRS. Cases and controls were subdivided into rare variant carriers and non-carriers. Risk for ALS was significantly associated with PRS and rare variants independently, but the interaction was not significant. ALS PRS affected risk only in those not carrying a rare variant, suggesting that rare variants in ALS genes are generally sufficient for disease risk. Rather than modifying the penetrance of rare variants, ALS PRS is most informative in the absence of these variants.

## INTRODUCTION

Amyotrophic lateral sclerosis (ALS) is a genetically heterogeneous neurodegenerative disease. Several biological pathways have been identified by which genetic variants can affect the pathogenesis of ALS^1^. However, phenotype is difficult to predict from single risk variants, and factors such as environmental exposures, occupation, ethnicity, and family history can modify the underlying risk, severity and onset of disease^2^.

Genetic analyses in ALS have been conducted in cohorts collected from ethnically diverse populations. These studies can highlight the unique genetic background of ALS in a given population; they can also help show how the genetic causes of a disease can be similar. There are several recent examples of population-specific examinations of ALS-associated genetic variants. In screening known ALS genes for variants in familial ALS cases from Germany, Muller et al. demonstrated phenotype/genotype correlations with previously reported as well as described novel variants in known genes^3^. A Turkish ALS population study found that 45% of familial ALS and 10% of sporadic ALS patients carried a variant in a previously reported gene; somewhat less than other European population screenings, indicating that differences between relatively close geographic regions can impart substantial effects on panel screening utility^4^. For example, in this Turkish cohort, expansion in *C9orf72* was the most common observed variant, but at a rate lower than expected based on Northern European prevalence estimates. Screening ALS genes originating from European studies becomes less practical in cohorts from Southern Africa^5^ or Japan^6^, for example, as fewer ALS cases may be genetically explained, or common genetic causes may be completely absent from the population (*C9orf72* expansion, notably). As unique genetic risk factors may occur in populations with diverse genetic ancestries, the identification of ALS genes and associated variants within them across a wide variety of populations is important if medical genetics is to be equitable for all individuals^7^.

The French-Canadian (FC) population of Québec has long been a focus of study in genetic epidemiology. Geographic and sociological factors throughout the history of Québec have resulted in a “founder effect,” by which certain alleles have been propagated within the population^8^. Multidimensional scaling of individual FC genetic variation has suggested that the geographic migration history of the region can still be captured by shared variants in the population^9^. Rare variants observed at a higher than expected frequency in the FC population have been observed to cause certain diseases^8^; some examples include autosomal recessive spastic ataxia of Charlevoix-Saguenay^10^, deafblindness from Usher syndrome^11^, and variants in the LDL receptor leading to familial hypercholesterolemia^12^. Rare variants found exclusively or at higher frequencies in the FC population could help better define ALS genetic risks.

In addition to rare ALS variants, the study of common variants can describe variability in disease risk. Several genome-wide association studies (GWAS) have been performed in ALS, with varying estimates explaining association and heritability^13-16^. Polygenic risk score (PRS) analysis is an approach to condense the association values obtained from GWAS into a per-sample score. Each variant has an associated level of risk or protection with the disease outcome, and variants can be summed into a PRS by the number of alleles and their quantitative weights (effect size). PRS for traits possibly related to ALS, such as schizophrenia^17^, epilepsy^18^, multiple sclerosis^19^, or blood lipids^20^ have had suggested associations, but few studies have examined the specificities of polygenic risk for ALS.

The intersection of rare and common variants can be informative for overall disease risk. Patients with low PRS could be considered a priority for rare variant screening^21^. Conversely, an individual without a rare, monogenic variant could have their disease partially explained by a high PRS^22^. Rare variants are often studied in terms of their combined burden across a gene; adding PRS to the statistical model can improve the fidelity of association by accounting for the inherited genetic liability^23,24^. In some diseases, such as hypertriglyceridemia, a high percentile of PRS can be equally or more predictive of disease outcome than the presence of a disease-associated rare variant^25^. In the context of ALS, this reasoning leads us to question if rare variants require common variants to be sufficiently pathogenic, and if polygenic risk is important in the context of penetrant rare variants.

In the current study, we aimed to describe the genetic architecture of ALS in the Québec FC population while testing the risk contributions of rare and common variants. We hypothesized that rare variants would be sufficient to impart risk for ALS without requiring increased PRS. We used targeted next-generation sequencing of known genes associated with ALS (as of the creation of the panel) and genome-wide SNP-chip genotyping to interrogate the contributions of both rare variants in coding regions with evidence of ALS risk and common variants across the genome, respectively. Our results characterize the specific genetics of ALS in the cohort, the distribution of rare variants in ALS genes of which several may be unique to the population, and the interplay between polygenic risk for ALS and rare risk variants in ALS genes.

## RESULTS

### Population structure of the FC genome

Following quality control (QC), relatedness, and genetic ancestry assessment, 327 cases and 344 controls remained for analysis. A total of 488,270 SNPs were included as input for imputation, and 29,359,578 SNPs remained after imputation QC. Principal component analysis (PCA) with 1000 Genomes Project (1KGP) data was performed using 78,170 common (MAF > 0.05) overlapping SNPs between cohort and 1KGP datasets. PCA plots showed a majority of the cohort overlapped with European individuals from 1KGP (Fig 1A-C). The contribution of different genetic backgrounds was shown in Admixture graphs (Fig 1D), with predominant proportions matching that of Northern and Western European (CEU) and British from England or Scotland (GBR) reference populations. A majority of the cohort was observed to have fewer than two genetic backgrounds from the admixture analysis, suggesting substantial genetic homogeneity.

**Figure 1:**
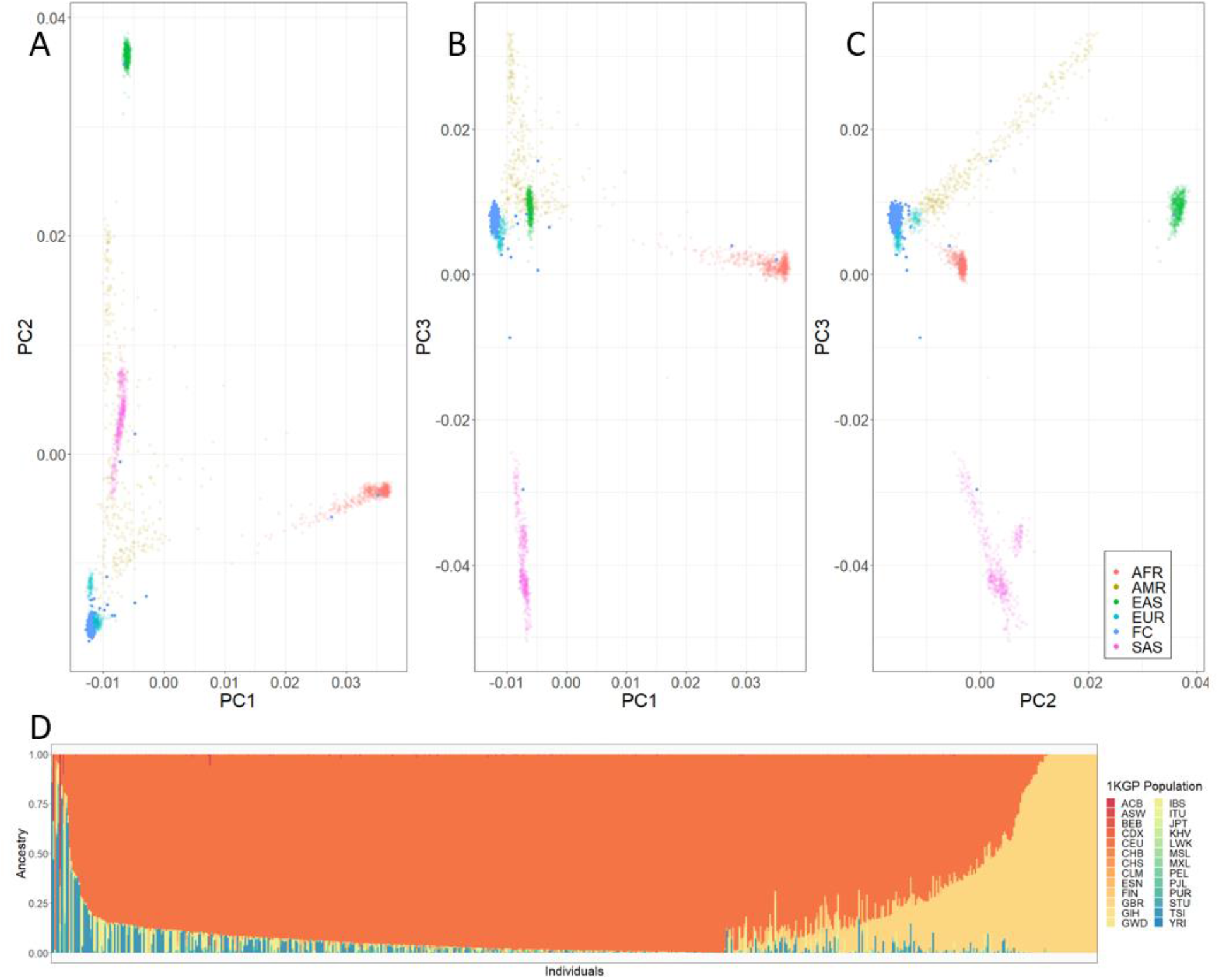
Common variant assessment of the Québec FC ALS cohort. A-C) Principal component analysis of common (minor allele frequency > 0.05) in the Québec ALS and unaffected control cohort compared to the 1000 Genomes Project (1KGP) reference populations. Samples in the Québec cohort cluster near the European super-population of the 1KGP cohort across the first three combinations of principal components (A: PC1 vs PC2, B: PC1 vs PC3, C: PC2 vs PC3). D) Admixture analysis of the Québec FC cohort using the 26 1KGP populations as references in a supervised analysis. Each vertical line of the stacked barplot represents a single cohort sample, with colours showing estimated reference population contributions.

### Rare variants are found throughout ALS genes

Following QC of sequencing data, 295 cases and 330 controls remained to test for rare protein-altering variants. 105 variants were observed across all individuals tested (Table 1), 23 of which had not previously been observed in the gnomAD Non-Finnish European (NFE) population^26^. Rare variants were observed in 36.3% of cases and 11.1% of controls (Figure 2). The *C9orf72* expansion was detected in 55 ALS cases (18.6%) and no controls.

**Table 1:**
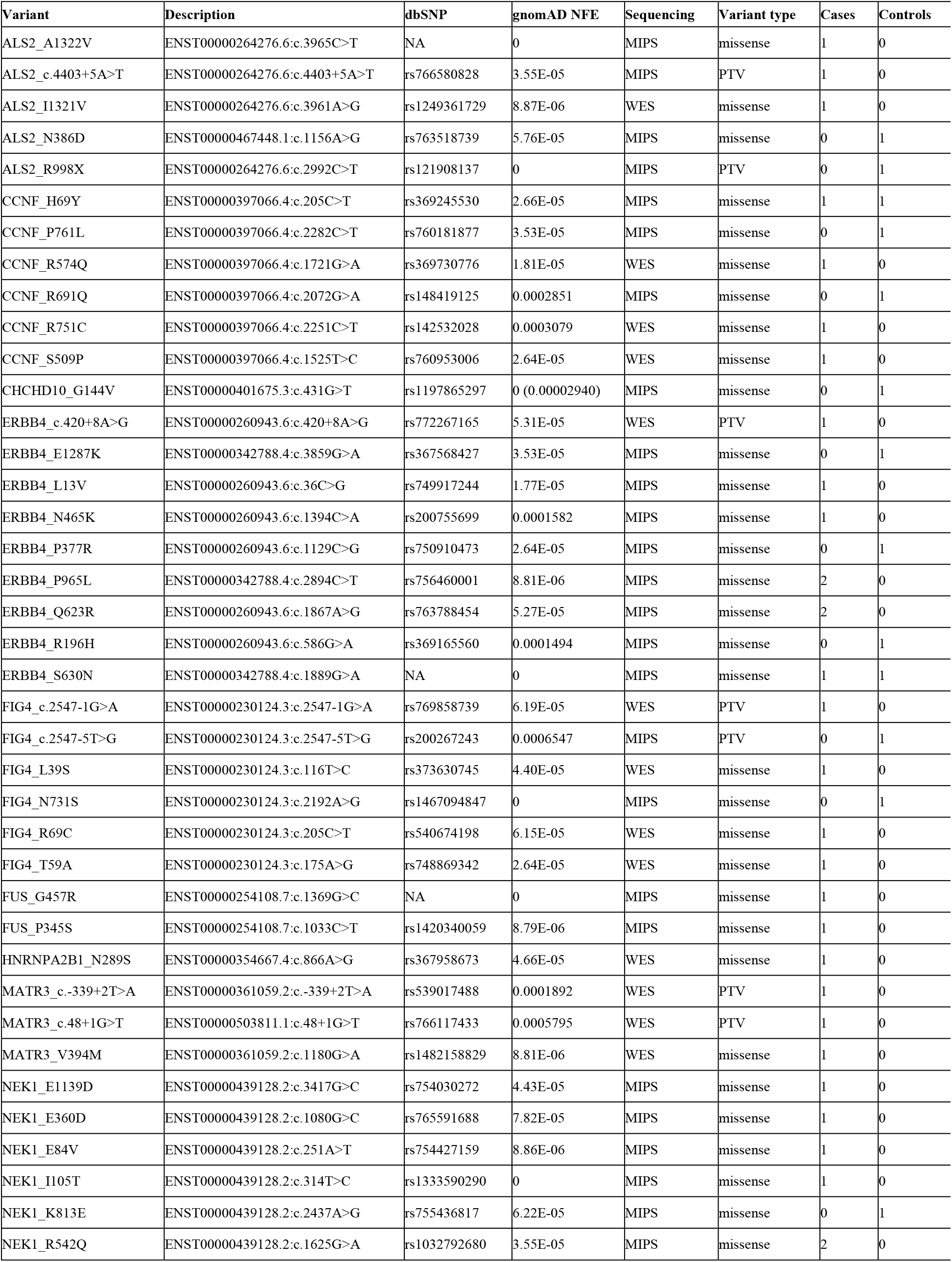
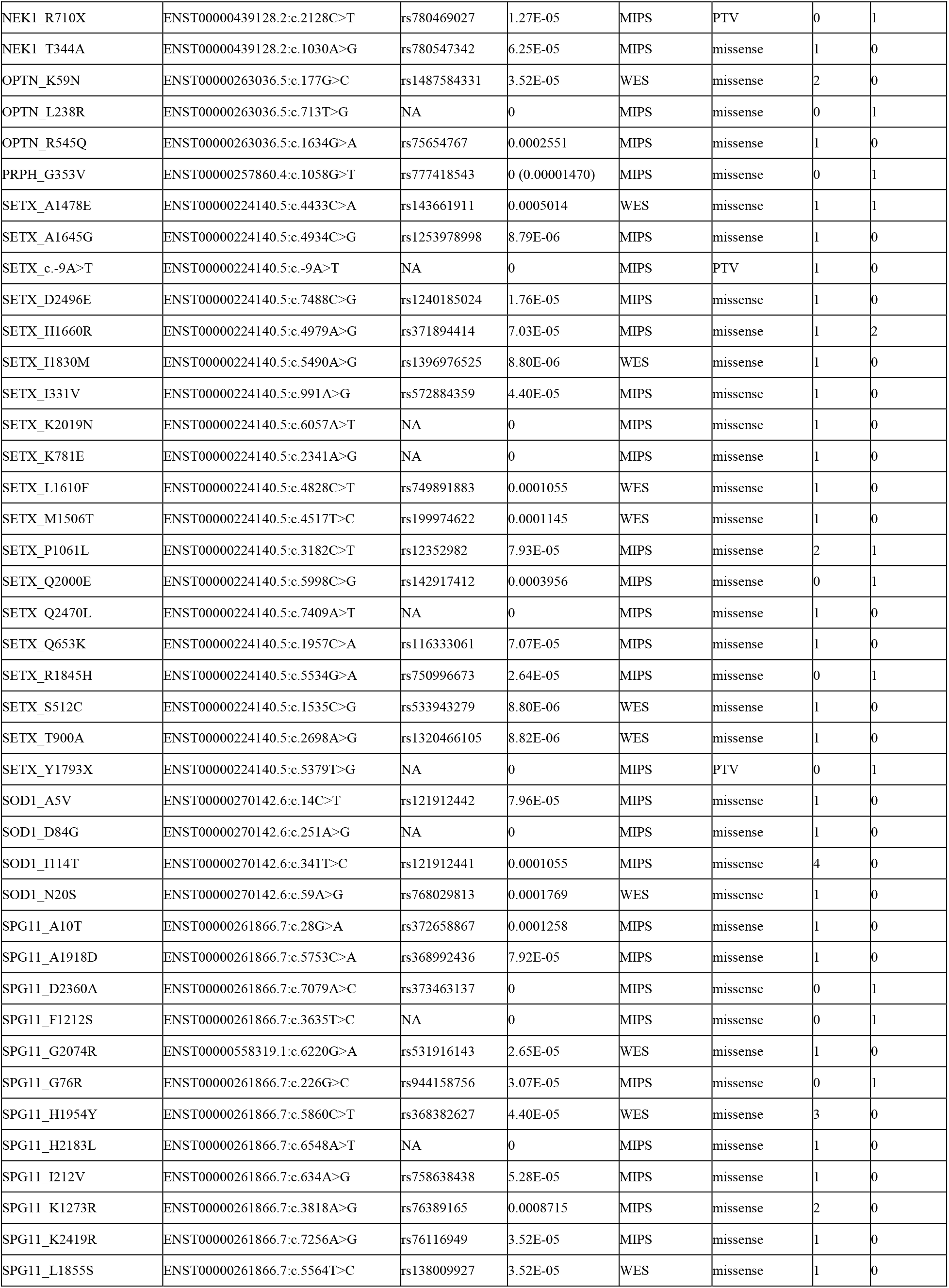
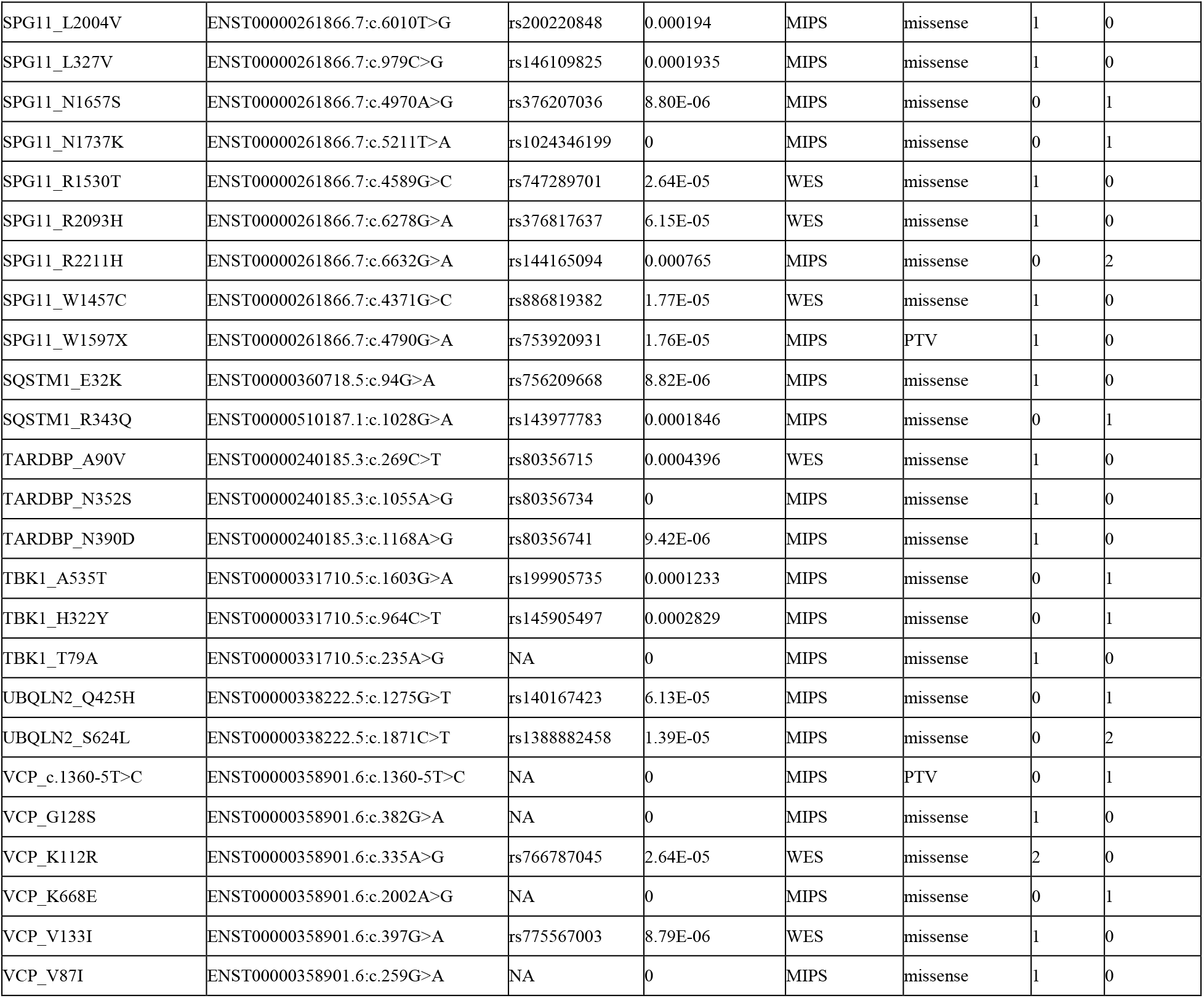
Rare variants observed in the Québec FC cohort. Variants are listed as Gene_Position. Description = Detailed description in the recommended format of Sequence Variant Nomenclature (HGVSC). dbSNP = existing variation by rsID in dbSNP. gnomAD_NFE = frequency of variant in the Non-Finnish European population of the gnomAD database v.2.1.1. Values in parentheses are updated frequencies for the variant in the updated gnomAD v.3.1.2 database. Sequencing = Type of sequencing in which variant is observed, targeted (MIPS) or exome sequencing (ES). Variant_type = consequence of variant on protein sequence, either protein truncating variant (PTV) or missense. Case and control counts are the number of individuals carrying the variant.

| Variant | Description | dbSNP | gnomAD NFE | Sequencing | Variant type | Cases | Controls |
| --- | --- | --- | --- | --- | --- | --- | --- |
| ALS2_A1322V | ENST00000264276.6:c.3965C>T | NA | 0 | MIPS | missense | 1 | 0 |
| ALS2_c.4403+5A>T | ENST00000264276.6:c.4403+5A>T | rs766580828 | 3.55E-05 | MIPS | PTV | 1 | 0 |
| ALS2_I1321V | ENST00000264276.6:c.3961A>G | rs1249361729 | 8.87E-06 | WES | missense | 1 | 0 |
| ALS2_N386D | ENST00000467448.1:c.1156A>G | rs763518739 | 5.76E-05 | MIPS | missense | 0 | 1 |
| ALS2_R998X | ENST00000264276.6:c.2992C>T | rs121908137 | 0 | MIPS | PTV | 0 | 1 |
| CCNF_H69Y | ENST00000397066.4:c.205C>T | rs369245530 | 2.66E-05 | MIPS | missense | 1 | 1 |
| CCNF_P761L | ENST00000397066.4:c.2282C>T | rs760181877 | 3.53E-05 | MIPS | missense | 0 | 1 |
| CCNF_R574Q | ENST00000397066.4:c.1721G>A | rs369730776 | 1.81E-05 | WES | missense | 1 | 0 |
| CCNF_R691Q | ENST00000397066.4:c.2072G>A | rs148419125 | 0.0002851 | MIPS | missense | 0 | 1 |
| CCNF_R751C | ENST00000397066.4:c.2251C>T | rs142532028 | 0.0003079 | WES | missense | 1 | 0 |
| CCNF_S509P | ENST00000397066.4:c.1525T>C | rs760953006 | 2.64E-05 | WES | missense | 1 | 0 |
| CHCHD10_G144V | ENST00000401675.3:c.431G>T | rs1197865297 | 0 (0.00002940) | MIPS | missense | 0 | 1 |
| ERBB4_c.420+8A>G | ENST00000260943.6:c.420+8A>G | rs772267165 | 5.31E-05 | WES | PTV | 1 | 0 |
| ERBB4_E1287K | ENST00000342788.4:c.3859G>A | rs367568427 | 3.53E-05 | MIPS | missense | 0 | 1 |
| ERBB4_L13V | ENST00000260943.6:c.36C>G | rs749917244 | 1.77E-05 | MIPS | missense | 1 | 0 |
| ERBB4_N465K | ENST00000260943.6:c.1394C>A | rs200755699 | 0.0001582 | MIPS | missense | 1 | 0 |
| ERBB4_P377R | ENST00000260943.6:c.1129C>G | rs750910473 | 2.64E-05 | MIPS | missense | 0 | 1 |
| ERBB4_P965L | ENST00000342788.4:c.2894C>T | rs756460001 | 8.81E-06 | MIPS | missense | 2 | 0 |
| ERBB4_Q623R | ENST00000260943.6:c.1867A>G | rs763788454 | 5.27E-05 | MIPS | missense | 2 | 0 |
| ERBB4_R196H | ENST00000260943.6:c.586G>A | rs369165560 | 0.0001494 | MIPS | missense | 0 | 1 |
| ERBB4_S630N | ENST00000342788.4:c.1889G>A | NA | 0 | MIPS | missense | 1 | 1 |
| FIG4_c.2547-1G>A | ENST00000230124.3:c.2547-1G>A | rs769858739 | 6.19E-05 | WES | PTV | 1 | 0 |
| FIG4_c.2547-5T>G | ENST00000230124.3:c.2547-5T>G | rs200267243 | 0.0006547 | MIPS | PTV | 0 | 1 |
| FIG4_L39S | ENST00000230124.3:c.116T>C | rs373630745 | 4.40E-05 | WES | missense | 1 | 0 |
| FIG4_N731S | ENST00000230124.3:c.2192A>G | rs1467094847 | 0 | MIPS | missense | 0 | 1 |
| FIG4_R69C | ENST00000230124.3:c.205C>T | rs540674198 | 6.15E-05 | WES | missense | 1 | 0 |
| FIG4_T59A | ENST00000230124.3:c.175A>G | rs748869342 | 2.64E-05 | WES | missense | 1 | 0 |
| FUS_G457R | ENST00000254108.7:c.1369G>C | NA | 0 | MIPS | missense | 1 | 0 |
| FUS_P345S | ENST00000254108.7:c.1033C>T | rs1420340059 | 8.79E-06 | MIPS | missense | 1 | 0 |
| HNRNPA2B1_N289S | ENST00000354667.4:c.866A>G | rs367958673 | 4.66E-05 | WES | missense | 1 | 0 |
| MATR3_c.-339+2T>A | ENST00000361059.2:c.-339+2T>A | rs539017488 | 0.0001892 | WES | PTV | 1 | 0 |
| MATR3_c.48+1G>T | ENST00000503811.1:c.48+1G>T | rs766117433 | 0.0005795 | WES | PTV | 1 | 0 |
| MATR3_V394M | ENST00000361059.2:c.1180G>A | rs1482158829 | 8.81E-06 | WES | missense | 1 | 0 |
| NEK1_E1139D | ENST00000439128.2:c.3417G>C | rs754030272 | 4.43E-05 | MIPS | missense | 1 | 0 |
| NEK1_E360D | ENST00000439128.2:c.1080G>C | rs765591688 | 7.82E-05 | MIPS | missense | 1 | 0 |
| NEK1_E84V | ENST00000439128.2:c.251A>T | rs754427159 | 8.86E-06 | MIPS | missense | 1 | 0 |
| NEK1_I105T | ENST00000439128.2:c.314T>C | rs1333590290 | 0 | MIPS | missense | 1 | 0 |
| NEK1_K813E | ENST00000439128.2:c.2437A>G | rs755436817 | 6.22E-05 | MIPS | missense | 0 | 1 |
| NEK1_R542Q | ENST00000439128.2:c.1625G>A | rs1032792680 | 3.55E-05 | MIPS | missense | 2 | 0 |
| NEK1_R710X | ENST00000439128.2:c.2128C>T | rs780469027 | 1.27E-05 | MIPS | PTV | 0 | 1 |
| NEK1_T344A | ENST00000439128.2:c.1030A>G | rs780547342 | 6.25E-05 | MIPS | missense | 1 | 0 |
| OPTN_K59N | ENST00000263036.5:c.177G>C | rs1487584331 | 3.52E-05 | WES | missense | 2 | 0 |
| OPTN_L238R | ENST00000263036.5:c.713T>G | NA | 0 | MIPS | missense | 0 | 1 |
| OPTN_R545Q | ENST00000263036.5:c.1634G>A | rs75654767 | 0.0002551 | MIPS | missense | 1 | 0 |
| PRPH_G353V | ENST00000257860.4:c.1058G>T | rs777418543 | 0 (0.00001470) | MIPS | missense | 0 | 1 |
| SETX_A1478E | ENST00000224140.5:c.4433C>A | rs143661911 | 0.0005014 | WES | missense | 1 | 1 |
| SETX_A1645G | ENST00000224140.5:c.4934C>G | rs1253978998 | 8.79E-06 | MIPS | missense | 1 | 0 |
| SETX_c.-9A>T | ENST00000224140.5:c.-9A>T | NA | 0 | MIPS | PTV | 1 | 0 |
| SETX_D2496E | ENST00000224140.5:c.7488C>G | rs1240185024 | 1.76E-05 | MIPS | missense | 1 | 0 |
| SETX_H1660R | ENST00000224140.5:c.4979A>G | rs371894414 | 7.03E-05 | MIPS | missense | 1 | 2 |
| SETX_I1830M | ENST00000224140.5:c.5490A>G | rs1396976525 | 8.80E-06 | WES | missense | 1 | 0 |
| SETX_I331V | ENST00000224140.5:c.991A>G | rs572884359 | 4.40E-05 | MIPS | missense | 1 | 0 |
| SETX_K2019N | ENST00000224140.5:c.6057A>T | NA | 0 | MIPS | missense | 1 | 0 |
| SETX_K781E | ENST00000224140.5:c.2341A>G | NA | 0 | MIPS | missense | 1 | 0 |
| SETX_L1610F | ENST00000224140.5:c.4828C>T | rs749891883 | 0.0001055 | WES | missense | 1 | 0 |
| SETX_M1506T | ENST00000224140.5:c.4517T>C | rs199974622 | 0.0001145 | WES | missense | 1 | 0 |
| SETX_P1061L | ENST00000224140.5:c.3182C>T | rs12352982 | 7.93E-05 | MIPS | missense | 2 | 1 |
| SETX_Q2000E | ENST00000224140.5:c.5998C>G | rs142917412 | 0.0003956 | MIPS | missense | 0 | 1 |
| SETX_Q2470L | ENST00000224140.5:c.7409A>T | NA | 0 | MIPS | missense | 1 | 0 |
| SETX_Q653K | ENST00000224140.5:c.1957C>A | rs116333061 | 7.07E-05 | MIPS | missense | 1 | 0 |
| SETX_R1845H | ENST00000224140.5:c.5534G>A | rs750996673 | 2.64E-05 | MIPS | missense | 0 | 1 |
| SETX_S512C | ENST00000224140.5:c.1535C>G | rs533943279 | 8.80E-06 | WES | missense | 1 | 0 |
| SETX_T900A | ENST00000224140.5:c.2698A>G | rs1320466105 | 8.82E-06 | WES | missense | 1 | 0 |
| SETX_Y1793X | ENST00000224140.5:c.5379T>G | NA | 0 | MIPS | PTV | 0 | 1 |
| SOD1_A5V | ENST00000270142.6:c.14C>T | rs121912442 | 7.96E-05 | MIPS | missense | 1 | 0 |
| SOD1_D84G | ENST00000270142.6:c.251A>G | NA | 0 | MIPS | missense | 1 | 0 |
| SOD1_I114T | ENST00000270142.6:c.341T>C | rs121912441 | 0.0001055 | MIPS | missense | 4 | 0 |
| SOD1_N20S | ENST00000270142.6:c.59A>G | rs768029813 | 0.0001769 | WES | missense | 1 | 0 |
| SPG11_A10T | ENST00000261866.7:c.28G>A | rs372658867 | 0.0001258 | MIPS | missense | 1 | 0 |
| SPG11_A1918D | ENST00000261866.7:c.5753C>A | rs368992436 | 7.92E-05 | MIPS | missense | 1 | 0 |
| SPG11_D2360A | ENST00000261866.7:c.7079A>C | rs373463137 | 0 | MIPS | missense | 0 | 1 |
| SPG11_F1212S | ENST00000261866.7:c.3635T>C | NA | 0 | MIPS | missense | 0 | 1 |
| SPG11_G2074R | ENST00000558319.1:c.6220G>A | rs531916143 | 2.65E-05 | WES | missense | 1 | 0 |
| SPG11_G76R | ENST00000261866.7:c.226G>C | rs944158756 | 3.07E-05 | MIPS | missense | 0 | 1 |
| SPG11_H1954Y | ENST00000261866.7:c.5860C>T | rs368382627 | 4.40E-05 | WES | missense | 3 | 0 |
| SPG11_H2183L | ENST00000261866.7:c.6548A>T | NA | 0 | MIPS | missense | 1 | 0 |
| SPG11_I212V | ENST00000261866.7:c.634A>G | rs758638438 | 5.28E-05 | MIPS | missense | 1 | 0 |
| SPG11_K1273R | ENST00000261866.7:c.3818A>G | rs76389165 | 0.0008715 | MIPS | missense | 2 | 0 |
| SPG11_K2419R | ENST00000261866.7:c.7256A>G | rs76116949 | 3.52E-05 | MIPS | missense | 1 | 0 |
| SPG11_L1855S | ENST00000261866.7:c.5564T>C | rs138009927 | 3.52E-05 | WES | missense | 1 | 0 |
| SPG11_L2004V | ENST00000261866.7:c.6010T>G | rs200220848 | 0.000194 | MIPS | missense | 1 | 0 |
| SPG11_L327V | ENST00000261866.7:c.979C>G | rs146109825 | 0.0001935 | MIPS | missense | 1 | 0 |
| SPG11_N1657S | ENST00000261866.7:c.4970A>G | rs376207036 | 8.80E-06 | MIPS | missense | 0 | 1 |
| SPG11_N1737K | ENST00000261866.7:c.5211T>A | rs1024346199 | 0 | MIPS | missense | 0 | 1 |
| SPG11_R1530T | ENST00000261866.7:c.4589G>C | rs747289701 | 2.64E-05 | WES | missense | 1 | 0 |
| SPG11_R2093H | ENST00000261866.7:c.6278G>A | rs376817637 | 6.15E-05 | WES | missense | 1 | 0 |
| SPG11_R2211H | ENST00000261866.7:c.6632G>A | rs144165094 | 0.000765 | MIPS | missense | 0 | 2 |
| SPG11_W1457C | ENST00000261866.7:c.4371G>C | rs886819382 | 1.77E-05 | WES | missense | 1 | 0 |
| SPG11_W1597X | ENST00000261866.7:c.4790G>A | rs753920931 | 1.76E-05 | MIPS | PTV | 1 | 0 |
| SQSTM1_E32K | ENST00000360718.5:c.94G>A | rs756209668 | 8.82E-06 | MIPS | missense | 1 | 0 |
| SQSTM1_R343Q | ENST00000510187.1:c.1028G>A | rs143977783 | 0.0001846 | MIPS | missense | 0 | 1 |
| TARDBP_A90V | ENST00000240185.3:c.269C>T | rs80356715 | 0.0004396 | WES | missense | 1 | 0 |
| TARDBP_N352S | ENST00000240185.3:c.1055A>G | rs80356734 | 0 | MIPS | missense | 1 | 0 |
| TARDBP_N390D | ENST00000240185.3:c.1168A>G | rs80356741 | 9.42E-06 | MIPS | missense | 1 | 0 |
| TBK1_A535T | ENST00000331710.5:c.1603G>A | rs199905735 | 0.0001233 | MIPS | missense | 0 | 1 |
| TBK1_H322Y | ENST00000331710.5:c.964C>T | rs145905497 | 0.0002829 | MIPS | missense | 0 | 1 |
| TBK1_T79A | ENST00000331710.5:c.235A>G | NA | 0 | MIPS | missense | 1 | 0 |
| UBQLN2_Q425H | ENST00000338222.5:c.1275G>T | rs140167423 | 6.13E-05 | MIPS | missense | 0 | 1 |
| UBQLN2_S624L | ENST00000338222.5:c.1871C>T | rs1388882458 | 1.39E-05 | MIPS | missense | 0 | 2 |
| VCP_c.1360-5T>C | ENST00000358901.6:c.1360-5T>C | NA | 0 | MIPS | PTV | 0 | 1 |
| VCP_G128S | ENST00000358901.6:c.382G>A | NA | 0 | MIPS | missense | 1 | 0 |
| VCP_K112R | ENST00000358901.6:c.335A>G | rs766787045 | 2.64E-05 | WES | missense | 2 | 0 |
| VCP_K668E | ENST00000358901.6:c.2002A>G | NA | 0 | MIPS | missense | 0 | 1 |
| VCP_V133I | ENST00000358901.6:c.397G>A | rs775567003 | 8.79E-06 | WES | missense | 1 | 0 |
| VCP_V87I | ENST00000358901.6:c.259G>A | NA | 0 | MIPS | missense | 1 | 0 |

**Figure 2:**
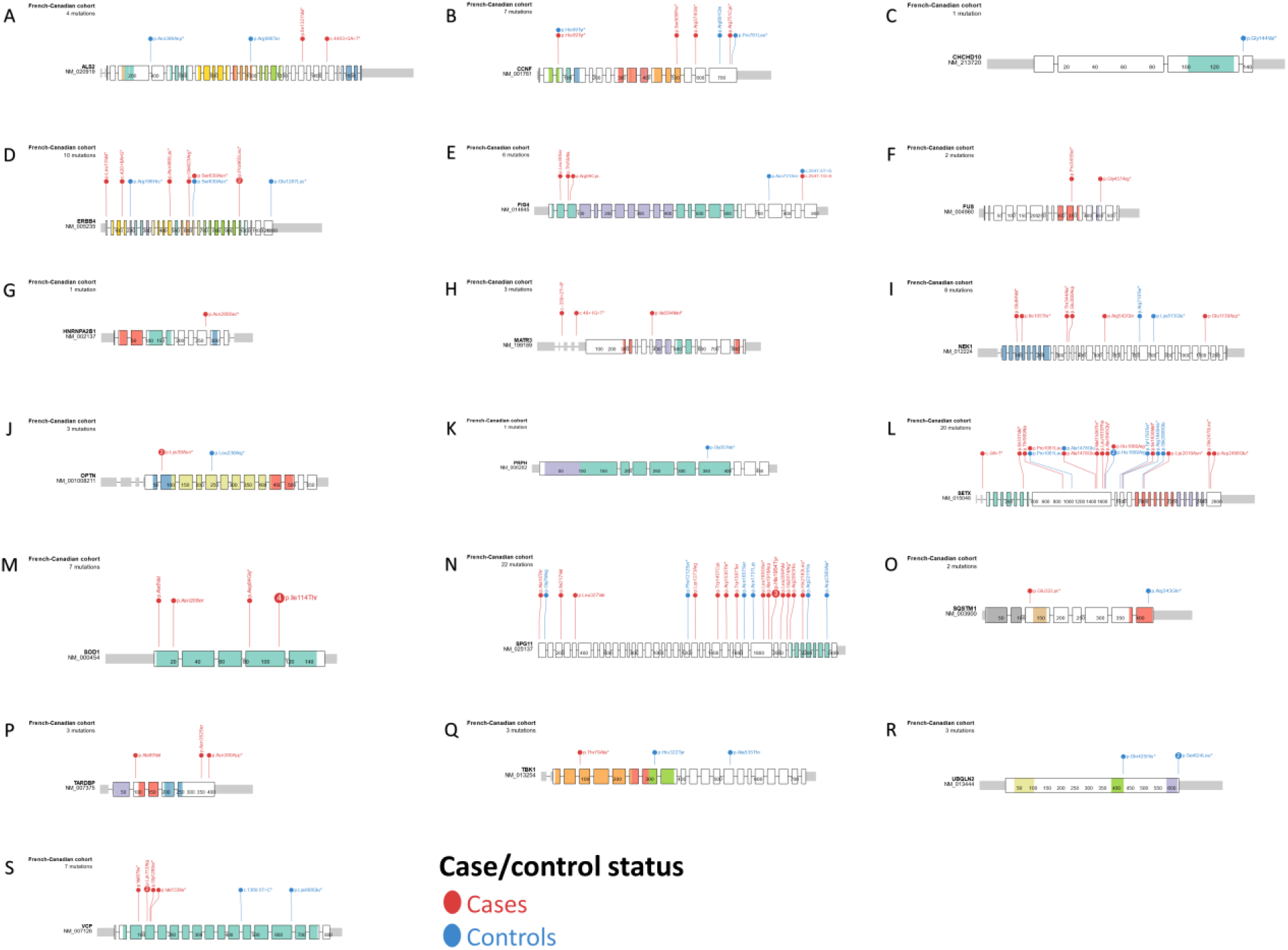
Rare variants observed in cases and controls of the FC ALS cohort. Gene profiles are shown with the locations of rare variants in ALS cases (red) or controls (blue). Exons are denoted by thicker boxes, while functional domains are indicated by coloured exons.

### Polygenic risk score explains slight but significant variability between ALS cases and unaffected controls

Following rare variant analysis, cases and controls were subsequently subset into carriers and non-carriers (Figure 3). After QC of genotyping, sequencing, and *C9orf72* results, 295 cases and 321 controls had sufficient data for both common and rare variant modalities and were included in PRS analysis. 6,635,680 variants were available from the ALS GWAS summary statistics, of which 5,251,483 variants overlapped with the imputed study dataset. After LD clumping using the 1KGP European samples^27^, 256,607 SNPs were used as input data for the PRS calculation. A PRS for ALS risk was generated for all cases and controls in our study cohort (model fit p = 8.21 × 10^−3^) (Figure 4A). This PRS explained 1.06% of variance between cases and controls, using a summary statistic threshold of p ≤ 2.95 × 10^−3^ and including 4,309 SNPs. The distribution of PRS in cases was significantly different from controls (t(607.52) = 3.13, p = 1.836 × 10^−3^) (Figure 4A), and individuals with PRS in the upper quartile (154 individuals per quartile) had a 1.92 odds ratio (95%CI = 1.14-3.26, p = 0.015) compared to individuals with PRS in the lowest quartile (Figure 4B). We assessed the distribution of SNPs across different functional genomic regions, and tested for biological process enrichment across the genes near which the SNPs were located (Figure 4C). We found that SNPs were enriched in genes that had ontology terms of cell migration, lipoprotein transport, and axon guidance.

**Figure 3:**
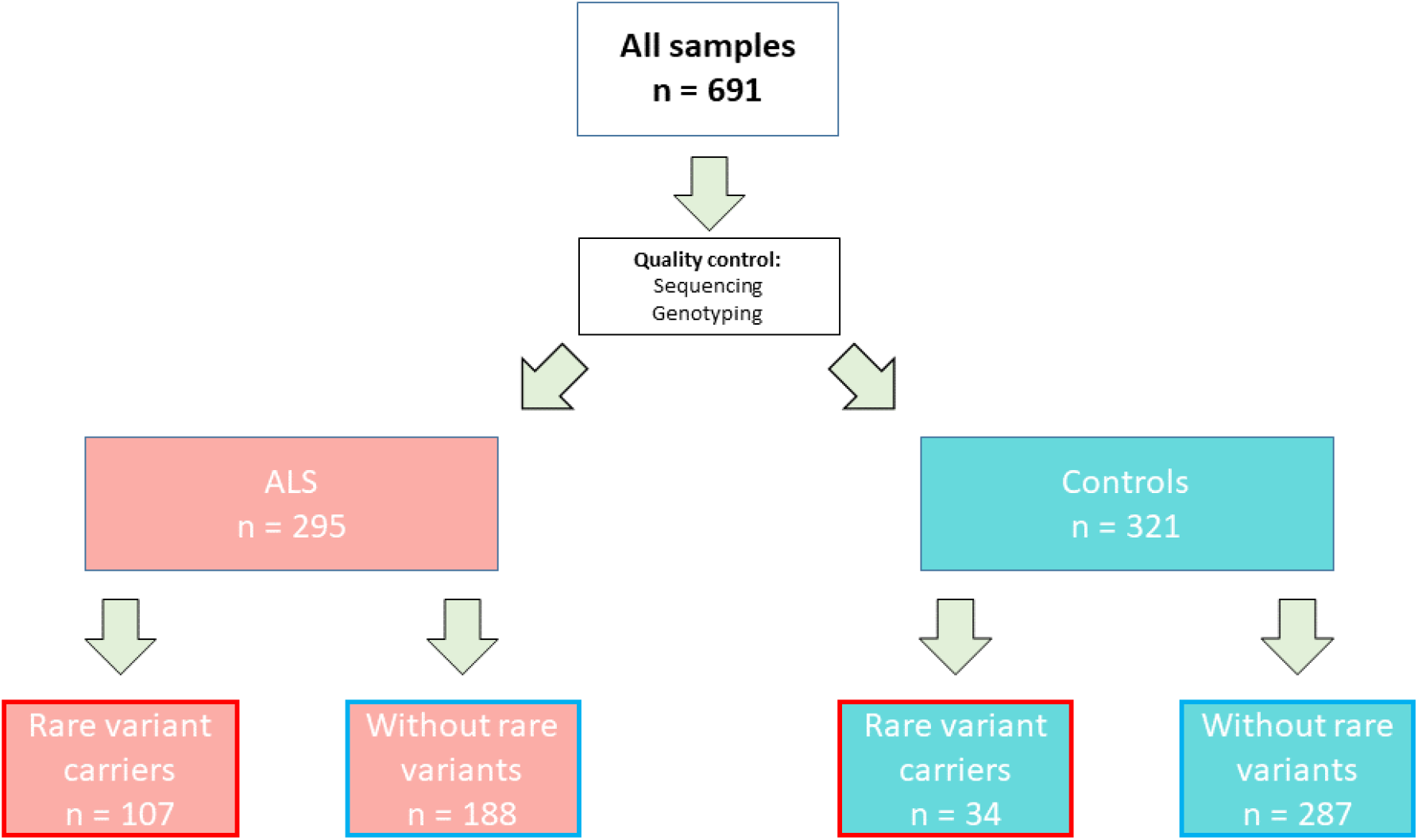
Flowchart of cohort sample types and division into rare variant subtypes. Sample numbers are given for the number of individuals included in the final overlapping PRS and rare variants analyses.

**Figure 4:**
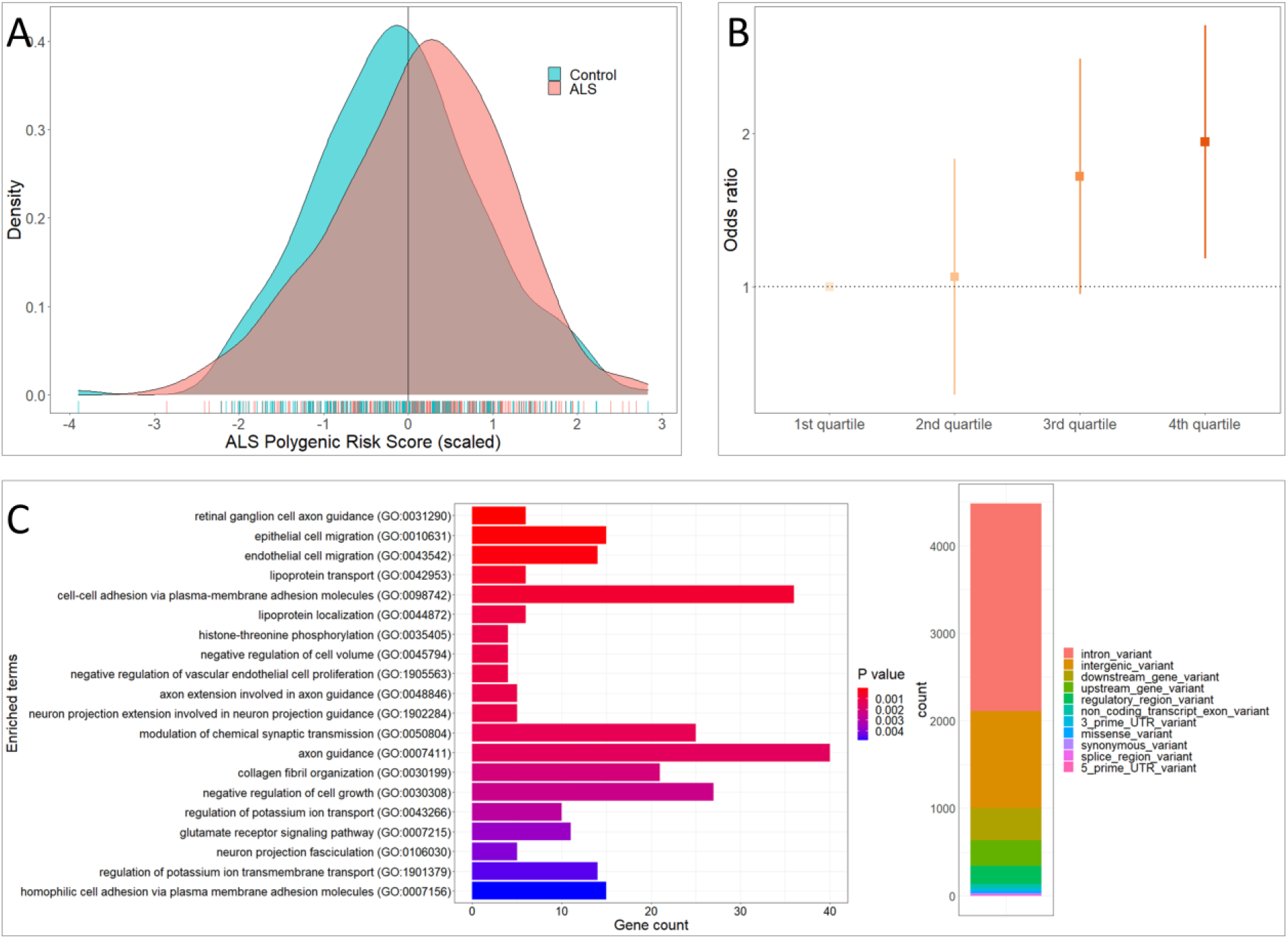
Polygenic risk score for ALS. A) Distribution of PRS between ALS cases and controls. PRS is shown as a Z-score scaled distribution, with the vertical line indicating the mean PRS in the cohort. Individual PRS are shown in the rugplot underneath the density graph. B) Odds ratios (OR) for each quartile (25%) of PRS distribution. Error bars represent the 95% confidence interval of the OR. OR values are relative to the first quartile as a baseline. 2nd quartile PRS OR = 1.05 (95%CI = 0.61-1.78, p = 0.87), 3rd quartile PRS OR = 1.65(95%CI = 0.97-2.80, p = 0.067), and 4th quartile PRS OR = 1.92 (95%CI = 1.14-3.26, p = 0.015) C) Distribution of SNP type and pathway enrichment of genes in which SNPs are nearest.

### Impact of polygenic risk for ALS depends on absence of rare variants

The predictive utility of PRS was different depending on the presence of rare variants. Logistic regression (including covariates of sex, genotyping batch, and the first 10 principal components (PC)) showed that both rare variant status and PRS were significantly associated with ALS outcome (OR = 5.10, 95%CI = 3.11-8.37, p = 1.06 × 10^−10^ for rare variants and OR = 1.35, 95%CI = 1.08-1.69, p = 9.96 × 10^−3^ for PRS), but not the interaction term of rare variants and PRS (OR = 0.871, 95%CI = 0.53-1.37, p = 0.551). ALS cases without a rare variant in ALS genes were the only subgroup to have a mean scaled PRS above the overall mean PRS (Figure 5A), suggesting that PRS is most informative in those cases without a known causal variant. Further, the OR for ALS of rare variant carriers was significantly above 1 for all quartiles of rare variant carriers, while only the third and fourth PRS quartiles of non-carriers had elevated OR compared to the reference of lowest quartile non-carriers (Figure 5B). Receiver operating characteristic (ROC) curve analysis showed that a model including an interaction between PRS and rare variant carrier status (including all previously mentioned covariates) was similarly predictive of ALS outcome as rare variants alone (AUC = 0.8363 and 0.8321, respectively), further supporting that rare variants and PRS contribute separately to ALS risk (Figure 5C). PRS varied between carriers with variants in different ALS genes, with a wide distribution across *C9orf72* expansion carriers (Figure 5D).

**Figure 5:**
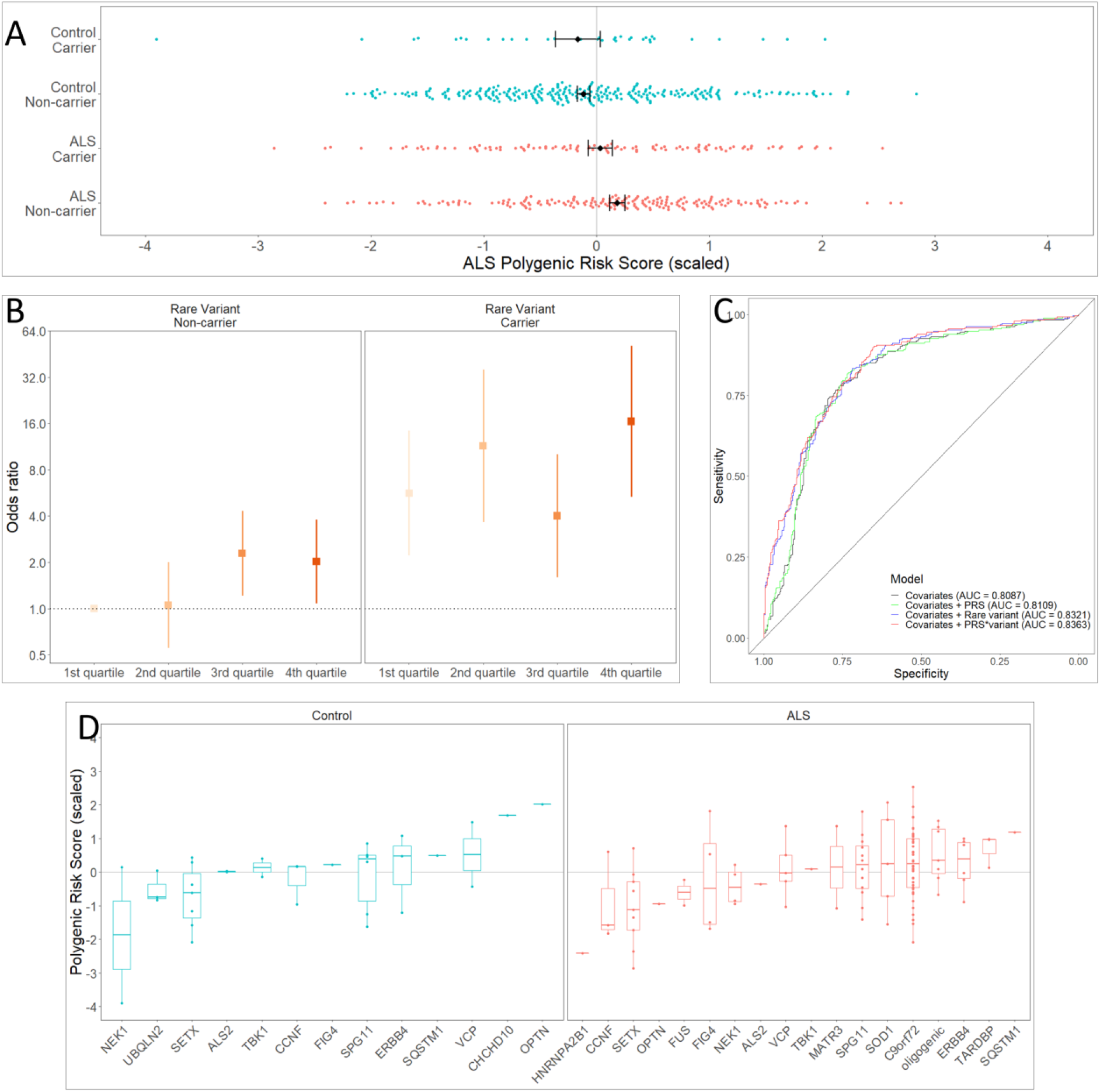
Polygenic risk score distributions by rare variant carrier status in ALS cases and controls. A) Distribution of PRS in each rare variant status subtype. PRS is Z-score scaled, with zero representing the cohort mean (grey vertical line). Black points overlaid on distributions represent the mean value of the subtype, and error bars represent the standard error of the mean. B) OR for each PRS quartile split by rare variant carrier status. Error bars represent the 95% confidence interval of the OR. OR values are relative to the first (lowest) quartile of the non-carrier subgroup. C) Receiver-operating characteristic (ROC) curves for models of PRS and ALS risk. Line colour represents different models incorporating various predictor variables, with area under the curve (AUC) listed for each. D) Distribution of PRS by gene in which a variant is observed. “Oligogenic” indicates that more than one variant in the same sample was observed.

## DISCUSSION

ALS has been studied as a monogenic or oligogenic disease, in which one or few concurrent rare variants are considered sufficient to instigate the disease^28^. However, the GWAS^14,16^ conducted for ALS have demonstrated substantial common variant associations. While the small effect sizes of most common variant associations likely do not lead to disease, inheriting several common variants may increase the overall disease risk. Our results suggest that this additive risk from common variants might play a role in risk for ALS in non-monogenic inheritance.

The PRS calculated from the current cohort showed a slight but significant difference in distribution between cases and controls. By subsetting the cohort into variant carriers and those without variants in ALS genes, the difference in PRS distribution was most apparent in those without rare variants. Since the PRS was not informative for variant carriers, the variants observed in these cases are likely sufficient to instigate disease *per se*. Conversely, the variants observed in controls may have been tolerated or coincidental. This interpretation aligns with the liability threshold (or multi-step) model of ALS, in which one or many genetic or environmental events are required to instigate disease. In this case, as the risk for ALS in rare variant carriers was not strongly affected by increased PRS, it could be viewed that the risk threshold is reached by rare variant alone, thus supporting sufficiency of rare variants in ALS genes.

ALS cases with a *C9orf72* expansion had a broad range of PRS. Currently it is difficult to assess large *C9orf72* expansion lengths, even with advancements in sequencing technology^29^. It is indeed likely that larger repeat lengths are more deleterious and penetrant than smaller lengths^30,31^, and future studies with accurate sizing data could evaluate the association between ALS PRS and *C9orf72* expansion length.

While the Québec FC population has been studied for founder effect variants, it does not appear that population-specific variants in known ALS genes play a large role in ALS risk. While we observed 23 variants that had not been reported in the gnomAD v2.1.1 NFE population, most of these variants were singletons, suggesting that variants in currently established ALS genes are likely not due to a founder effect. If such an effect plays a role in ALS, it could be due to genes that were not examined in this study, and could be yet unknown in ALS research.

Our study has several limitations. Our sample size was limited, and as such we might not have observed certain rare variants that had interactions with PRS. We observed that the interaction term between PRS and rare variants was not significant; however, it may be that only a certain type of variant interacts with PRS to modify ALS risk. Because exome sequencing was available only for cases, there may be an underestimation of variants in the control cohort, as ES is generally more sensitive to detect variants. We used the older gnomAD v2.1.1 as a variant frequency reference while v3.1.2 is currently available. The matched genomic assembly and increased ES use of v2.1.1 allowed a better comparison between our dataset and the gnomAD reference. We examined whether novel variants in our cohort were observed in the latest gnomAD version to account for this limitation. Finally, the PRS analysis used GWAS summary statistics from the 2016 ALS GWAS^14^. While there has recently been a more recent GWAS with a larger sample size^16^, the samples included in the current study were also included in the newest GWAS, and therefore overfitting of the PRS model would occur.

As increasingly monogenic, rare causes of ALS are uncovered, and other well-powered GWAS are performed, the contribution of genetics in ALS will be better understood. The interplay between rare and common variants might differ between genetic ancestries, as certain rare variants are ancestry-specific and common variants are affected by population drift and geographic distance. Future studies could also incorporate other types of variants, such as structural or copy-number, as new technologies allow for better sequencing accuracy. ALS remains a heterogeneous disease, and with deeper phenotyping from large cohorts, a more complete understanding of genetic impacts on disease onset and progression may be attained.

## METHODS

### Sample cohorts

In total, 335 ALS cases and 356 controls from clinics across Québec, Canada were included in this study. All samples were sequenced by either custom-design targeted panel, whole exome sequencing, and were tested for the *C9orf72* repeat expansion in order to detect rare variants in ALS genes. Samples with a *C9orf72* expansion were not subjected to further sequencing. Of these samples, 329 cases and 349 controls were genotyped by SNP-chip genotyping. Sample details are summarized in Table S1.

Cases were included agnostic of symptom presentation, family history, or presence of a disease-causing variant. Controls were included if they had no known relative with ALS, were at least 50 years old at time of sampling, and did not have a clinical diagnosis of a neurodegenerative or psychiatric disorder. Average ages of cases and controls across the cohort were 60.5 ± 12.2 years and 62.2 ± 9.0 years, respectively. Sex ratios (male:female) for cases and controls were 1.61 and 0.967, respectively. All individuals signed informed consent and samples were collected following approved ethics protocols from ethics review boards of all institutions involved. Blood DNA was extracted using standard salting out procedures.

### Genotyping

All genotyping was performed using the Global Screening Array version 2 (Illumina, Inc.) at the McGill University and Génome Québec Innovation Centre (MUGQIC). QC was performed using the default settings of the RICOPILI QC module (v2019_Oct_15.001)^32^. Briefly, the criteria were: ≥98% call rate for variants, ≥98% call rate for samples, ≤0.02 call rate difference between cases and controls, Hardy-Weinberg Equilibrium (HWE) p > 1×10^−6^ in controls and HWE p > 1×10^−10^ in cases, and inbreeding coefficient outside of ±0.2.

Samples were tested for relatedness using the KING algorithm in the SNPRelate^33^ R package, and pairwise exclusion of samples was performed for kinship above 0.125 (second degree relatives). Samples were limited to those of inferred European descent by comparison with the 1KGP superpopulations^27^ using PCA calculated using SNPRelate^33^. The first four PCs were used to exclude samples that did not overlap with samples of known European ancestry. PCA was then performed on the case control cohort without 1KGP data to calculate principal components for use as covariates. Admixture analysis was performed after QC using the software ADMIXTURE^34^. The 1KGP dataset was used to perform a supervised analysis, with each of the 26 populations set as a known ethnicity and the case-control cohort set as unknown.

Following QC, genotyping data was imputed on the Sanger Imputation Service using the Haplotype Reference Consortium (release 1.1) panel with EAGLE2 pre-phasing^35^. Imputed SNPs were filtered for imputation R^2^ > 0.3 and for HWE p > 1×10^−5^. The program BCFtools v.1.10 from the SAMtools software suite^36^ was used for both VCF imputation preparation and post-imputation filtering.

### Rare variant analysis

Sequencing was performed using targeted sequencing by Molecular Inversion Probe Sequencing (MIPS) or Exome Sequencing (ES). All sequencing was generated at the McGill University and Génome Québec Innovation Centre (MUGQIC). Sequencing data was aligned to the GRCh37 reference genome. ES variant calls were limited to the same genomic regions as the MIPS. MIPS and ES processing was performed similarly to previous studies^37,38^, with minor changes described here. For ES, the Genome Analysis ToolKit (GATK) v3.7^39,40^ was used to joint genotype samples into a combined dataset, while for MIPS VarScan v2.3.9 was used^41^. Variants called from GATK were required to have GQ ≥30, DP ≥20, and call rate ≥90%. Variants called from MIPS with VarScan were required to have minimum quality ≥20, minimum coverage ≥20, and minimum allele balance ≥0.25 for heterozygous calls. Samples were limited to those with ≥70X average sequencing depth and that had ≥90% call rate of post-QC variants in both MIPS and ES. Variants included following quality control were annotated for frequency and functional consequences using VEP version 98^42^. Variants were limited to those with a minor allele frequency ≤ 0.001 in the NFE from gnomAD version 2.1.1^26^ and that were predicted to be missense, stop-lost, stop-gained, or splice site altering. Samples were tested for *C9orf72* repeat expansion as previously described^43,44^. Samples that passed both genotyping and sequencing QC were assigned to a subtype of cases or controls (Figure 1). Variants were plotted on gene diagrams using PeCan (https://pecan.stjude.cloud/proteinpaint). All bioinformatic analyses were performed on the Béluga and Narval computing clusters of Compute Canada and Calcul Québec.

### Polygenic risk score calculation

ALS PRS was calculated with PRSice-2^45^, using the summary statistics from a meta-analysis of ALS-GWAS^14^. The first 10 PCs, sex, genotyping batch, and rare variant carrier status were included as covariates in the PRS calculation. Because age was used as a threshold for controls it was not included in the model. The 1KGP European cohort was used to account for linkage disequilibrium.

### Statistical analyses

All statistical analyses were performed using R v3.5.1. A Welch’s two-sample t-test was used to test for difference in mean PRS between cases and controls. The association between ALS outcome and PRS or rare variant status was assessed using a logistic regression. Covariates included in the model were the first 10 PCs, sex, genotyping batch, and the interaction between PRS and carrier status. Logistic regression was similarly used to assess the odds ratio of each intersection between PRS quartile and rare variant status. ROC curves were calculated using the R package pROC^46^ to test between model of ALS ∼ PRS or ALS ∼ PRS*carrier-status, including all covariates. Pathway enrichment analysis of the SNPs comprising the PRS was performed using EnrichR^47^. SNP functional consequences were annotated using vep (cache version 98)^42^.

## Data availability

Raw sequencing results will be available on the NCBI SRA database (accession to be determined after publication). Calculated PRS per sample are provided as raw values in Table S1. Genomic regions targeted by MIPS probes are listed in Table S2.

## Data Availability

Raw sequencing results will be available on the NCBI SRA database (accession to be determined after publication).

## ACKNOWLEDGEMENTS

We would like to thank the participants of the study. We thank Hélène Catoire and Vessela Zaharieva for assistance in clinical coordination and Cynthia V. Bourassa and Sandra Laurent for technical assistance. J.P.R. has received a doctoral student fellowship from the ALS Society of Canada and a Canadian Institutes of Health Research Frederick Banting & Charles Best Canada Graduate Scholarship (FRN 159279). We thank the ALS Society of Canada, Brain Canada, the Canadian Institutes of Health Research, and the Radala Foundation for ALS Research for research funding.

